# A Single-cell Atlas of Juvenile Nasopharyngeal Angiofibroma Reveals VEGF-Driven Angiogenic Remodeling as a Therapeutic Vulnerability

**DOI:** 10.64898/2026.07.01.26356778

**Authors:** Heidi Martini-Stoica, Brittany T. Rupp, Meik Kunz, Alessandra Livraghi-Butrico, Kenichi Okuda, Wanda K. O’Neal, Scott H. Randell, Hong Dang, Hiroaki Murano, Minako Furusho, Lisa Morton, Frederic Askin, Brian D. Thorp, Cristine Klatt-Cromwell, Charles S. Ebert, Brent A. Senior, Jackson R. Vuncannon, Adam J. Kimple, Kevin M. Byrd

## Abstract

**Background:** Juvenile nasopharyngeal angiofibroma (JNA) is a rare locally aggressive vascular sinonasal tumor that primarily affects adolescent males. Despite advances in endoscopic surgery and preoperative embolization, JNA can be associated with major operative bleeding risk and clinically meaningful recurrence, while non-surgical treatment options remain limited.

**Methods:** To define the cellular programs underlying JNA vascularity, we performed single-cell RNA sequencing of JNA tumors (n=2), tumor-adjacent mucosa, and control sinonasal tissue. We analyzed cell composition, differential gene expression, pathway enrichment, and cell–cell communication, followed by Drug2cell-based mapping of transcriptional states to candidate therapeutic targets.

**Results:** JNA contained an expanded fibrovascular compartment composed of endothelial cells, fibroblasts, pericytes, vascular smooth muscle cells, and neural crest–like cells. Neural crest–like cells were enriched in JNA but showed relatively limited transcriptional differences from tumor-adjacent tissue. By contrast, endothelial cells demonstrated the strongest disease-associated remodeling, with enrichment of angiogenesis, extracellular matrix organization, hypoxia response, and cell migration pathways. Endothelial cells also showed downregulation of adaptive immune signaling pathways, suggesting reduced immune engagement within the tumor microenvironment. Intercellular communication analyses revealed dense endothelial–stromal signaling across the JNA fibrovascular network. Drug2cell analysis nominated VEGF/VEGFR signaling as a candidate therapeutic vulnerability, with VEGFR-targeting agents predicted to act primarily on vascular and lymphatic endothelial populations.

**Conclusions:** JNA is organized around an angiogenesis-dominant fibrovascular program driven by endothelial-centered signaling. These data support further investigation of VEGF/VEGFR-directed therapy as a potential adjunctive strategy for patients with recurrent, unresectable, or surgically high-risk JNA.

## Introduction

Juvenile nasopharyngeal angiofibroma (JNA) is a rare, benign but locally aggressive vascular tumor of the sinonasal tract.^1^ JNA typically presents in adolescent male subjects, usually in the second decade of life.^2^ The tumor most commonly arises from the vicinity of the sphenopalatine foramen and demonstrates a locally aggressive pattern of growth with extension into adjacent structures, including the nasal cavity, nasopharynx, paranasal sinuses, orbit, and skull base, while typically displacing rather than invading normal tissues.^3^ Despite its benign histology, the anatomic location, marked vascularity, and potential for skull base extension make JNA clinically challenging.

The standard of care for treatment of JNA is surgical resection, via endoscopic or open approaches, typically following pre-operative embolization.^2,4^ Although surgical outcomes have improved, recurrence remains clinically meaningful, with recurrence rates following surgical resection range from 4 to 40%, impacted by factors such as tumor stage, intracranial extension, incomplete resection, and open versus endoscopic technique.^2,5,6^ Treatment of recurrent JNA includes repeat surgery if resectable, radiation therapy if unresectable, or observation.^3,4,7^ These clinical challenges highlight the need for improved biologic understanding and alternative therapeutic strategies for JNA.

Histopathologically, JNA is composed of a connective tissue stroma with extensive vasculature lined by a single layer of endothelial cells and occasional smooth muscle cells, but lacking a continuous muscular layer and stromal elastic fibers.^3,6,8^ These structurally abnormal vessels are thought to contribute to the tumor’s propensity for hemorrhage and reflect dysregulated vascular maturation. Debate persists as to whether JNA is a true tumor or a vascular malformation arising from remnants of the first branchial arch artery plexus, with some studies proposing neural crest cells as the cell of origin.^9^ This unresolved question has direct biologic relevance because neural crest–derived cells contribute to craniofacial vascular development and may help explain the characteristic location and vascular phenotype of JNA. Therefore, the cellular composition and functional organization of these vascular and stromal compartments require further investigation.

Angiogenic factors are a particular focus given the highly vascular nature of the tumor. Vascular endothelial growth factor (VEGF), a pro-angiogenic growth factor, as well as its receptors and downstream targets, have been generally found to be upregulated in JNA.^10–15^ In particular, high levels of VEGFR2 are linked to increased vessel density in JNA.^16^ Furthermore, Mishra et al demonstrated that increased mRNA levels of *VEGF* correlated with clinical features such as intraoperative hemorrhage, tumor volume, skull base extension, and recurrence.^17,18^ Additional pathways, including FGF, PDGF, NGF, and IGFs, have also been implicated in JNA.^10,14,17,18^ These studies support angiogenic signaling as a central feature of JNA biology. However, bulk tissue and immunohistochemical studies cannot fully determine which cell populations produce or respond to these signals, how these pathways are coordinated across the tumor microenvironment, or which cellular compartments may be most therapeutically targetable.

Therapeutic options in JNA beyond surgery and radiation are limited. Due to JNA presenting almost exclusively in male subjects around the time of puberty, the role of androgen receptor signaling has been investigated and its inhibition was initially hypothesized to have therapeutic potential.^3^ However, treatment with flutamide, an androgen receptor antagonist, provided inconsistent results with regard to reducing tumor volume.^19^ The only current clinical trial for JNA is evaluating the safety and efficacy of sirolimus, an mTOR inhibitor previously utilized with success in treating vascular malformations.^20–22^ However, these approaches are not specifically tailored to the underlying biology of JNA and may not fully address the mechanisms driving its vascular growth and recurrence. Additional targeted strategies, including anti-angiogenic therapies, have been proposed based on prior evidence of VEGF pathway activation, but have not been systematically evaluated in this disease. The absence of a detailed, cell type–resolved understanding of JNA limits the ability to deploy such therapies. Consequently, there remains a lack of targeted treatments informed by tumor-specific biology.

Given the limited availability of targeted therapies beyond surgery and radiation, we applied single-cell transcriptomic profiling to an exploratory cohort of JNA tumors, adjacent mucosa, and control sinonasal tissue to define the tumor microenvironment at cellular resolution. This approach enables identification of the cellular sources and target populations of angiogenic and stromal signaling, characterization of intercellular communication networks, and prioritization of candidate therapeutic vulnerabilities. By resolving the angiogenesis-dependent fibrovascular architecture of JNA, this study provides a framework for understanding its clinical behavior and for identifying therapeutic strategies for patients with recurrent or unresectable disease.

## Methods

### Patient recruitment and medical imaging

Patients were seen and recruited at the University of North Carolina Hospital. Patients with presumed juvenile nasopharyngeal angiofibroma (JNA) based on clinical history, nasal endoscopy, and CT/MRI imaging were included. Definitive diagnosis of JNA was later confirmed by surgical pathology. Healthy control patients were selected based on having no history of chronic sinus disease and were already scheduled for minimally invasive pituitary surgery. Informed consent was obtained from each patient and/or their legal guardian under an approved IRB (UNC #03-1396) and in accordance with the Declaration of Helsinki. Tumor and tumor adjacent tissues were obtained from JNA patients during surgical resection, while healthy control tissue was obtained from the ethmoid sinuses of patients undergoing surgery. Patient demographic information can be found in Table 1.

**Table 1:** Patient demographic information.

| ID | Sex | Age Range | Race | Ethnicity |
| --- | --- | --- | --- | --- |
| JNA 1 | Male | 11-15 | Caucasian | Non-Hispanic |
| JNA 2 | Male | 11-15 | Caucasian | Non-Hispanic |

Computed Tomography (CT) and Magnetic Resonance Imaging (MRI) were performed on JNA patients as part of the preoperative workup. All JNA patients underwent targeted embolization of tumor feeder vessels the day prior to resection.

### Sample collection and processing

Excess resected tissue samples that were not needed for diagnosis were collected and split for paraffin embedding and single cell RNA sequencing (scRNAseq). The portion for paraffin embedding was immediately fixed in 10% buffered formalin, while samples for scRNAseq were dissociated into a single-cell suspension as previously described.^23^

### Histology

Formalin-fixed, paraffin-embedded tissue blocks were sectioned, deparaffinized, rehydrated and stained using hematoxylin and eosin (H&E).

### Single-cell RNA sequencing (scRNA-seq)

Dissociated tissue samples were processed using the Chromium Controller (10x Genomics) to generate single-cell gel bead-in-emulsions (GEMs), and libraries were prepared using the Chromium Next GEM Single Cell 3′ Kit v3.1 (10x Genomics) according to the manufacturer’s instructions. Following reverse transcription and cDNA amplification, libraries were constructed and purified using SPRIselect reagent (Beckman Coulter, cat. #B23317). Sequencing libraries were quantified using TapeStation (Agilent) and sequenced on a Illumina NovaSeq X Plus (JNA tumor, tumor adjacent samples) and NextSeq2000P4 (healthy samples) using paired-end reads to achieve a minimum depth of 40,000 read pairs per cell.

### Data processing

FASTQ files generated from sequencing were processed using CellRanger with intron mode (10x Genomics) and mapped to the human reference genome (GRCh38.gc42.CR7). Processed feature.tsv, matrix.mtx, and barcode.tsv files were exported from CellRanger and uploaded to Trailmaker (Parse Bioscience) for additional filtering and data analysis. Within Trailmaker samples underwent the following filtering: False discovery rate of 0.01 for cells versus ambient RNA, minimum number of transcripts per cell equal to 500, maximum percentage of mitochondrial reads equal to 15%, and droplet probability threshold of 0.5.

After filtering, cell populations were annotated within Trailmaker using known gene expression profiles. Uniform Manifold Approximation and Projection (UMAP) embeddings, dot plots, and bar graphs were generated using the plots and tables feature. Five unique cell subpopulations (Vascular smooth muscle, Pericytes, Neural crest-like cells, fibroblasts, and endothelial cells) were subset and used to construct volcano plots comparing tumor and tumor-adjacent tissues. Differentially expressed genes (DEGs) were determined using cutoffs of FDR<0.05 and log_2_(fold change) >0.25. DEG lists were compiled for each subpopulation and analyzed by gene ontology analysis using DAVID.^24,25^

### Cell-cell communication analysis

Cell–cell communication was inferred from the single-cell transcriptomic dataset using CellPhoneDB v5.0.0 and CellChatv2 as we have done previously.^26–28^ Normalized gene expression matrices annotated with Ensembl gene identifiers were analyzed together with cell-level metadata defining cell-type assignments. Ligand–receptor interactions were evaluated across annotated cell populations using both non-statistical and permutation-based CellPhoneDB workflows, with interaction scoring enabled and genes retained when expressed in at least 10% of cells within a given cell type. Statistical significance was assessed using 1,000 random permutations, a fixed random seed for reproducibility, and a significance threshold of P < 0.05. To prioritize context-relevant interactions, differentially expressed genes were incorporated into a DEG-aware CellPhoneDB analysis, generating relevant interaction matrices and mean expression scores across interacting cell-type pairs. Downstream visualization and prioritization were performed using ktplotspy. CellChat (v2) quantified the signaling probabilities between sender and receiver cell groups. Resulting circle plots showed the number of interactions between annotated cell types within both tumor samples and adjacent tissue samples. The thickness of the lines within the circle plot represents the number of cell interactions, while the color matches the color of the cell type acting as the sender of the signal. Node size was proportional to the number of cells in each group.

### Drug2cell

Single-cell transcriptomic datasets were analyzed using drug2cell to infer cell-state-specific drug response signatures from expression profiles, as has been done previously.^29–31^ Processed AnnData objects were loaded into Scanpy, UMAP embeddings were harmonized for visualization, and drug2cell scores were computed using raw gene expression values. Drug-enrichment profiles were generated for the full dataset and for disease-relevant subsets. Differential drug signature enrichment across annotated cell populations and disease states was assessed using Wilcoxon rank-sum testing implemented in Scanpy’s rank_genes_groups function. Ranked drug signatures, log-fold changes, nominal P values, and adjusted P values were exported for downstream interpretation. Top-ranked drug associations were visualized as dot plots, including unbiased top-five summaries and curated drug-of-interest panels grouped by therapeutic class or category.

## Results

### Single-Cell Profiling Defines the Cellular Composition of JNA

Single-cell RNA sequencing (scRNA-seq) was performed on tumor and healthy tumor-adjacent sinonasal tissue from two adolescent males diagnosed with JNA, who underwent endoscopic surgical resection of their tumor. Representative CT and MRI imaging demonstrated the pathognomonic Holman-Miller antral sign, as well as enhancement with contrast and heterogeneous appearance on T2 weighted imaging due to flow voids (Figure 1a,b). Representative H&E staining of JNA tumor demonstrate a highly fibrovascular tumor (Figure 1c). Healthy control samples were obtained from ethmoid sinuses of adult patients undergoing minimally invasive pituitary surgery for comparison. No evidence of neoplastic involvement was present in control or tumor-adjacent tissues.

**Figure 1.**
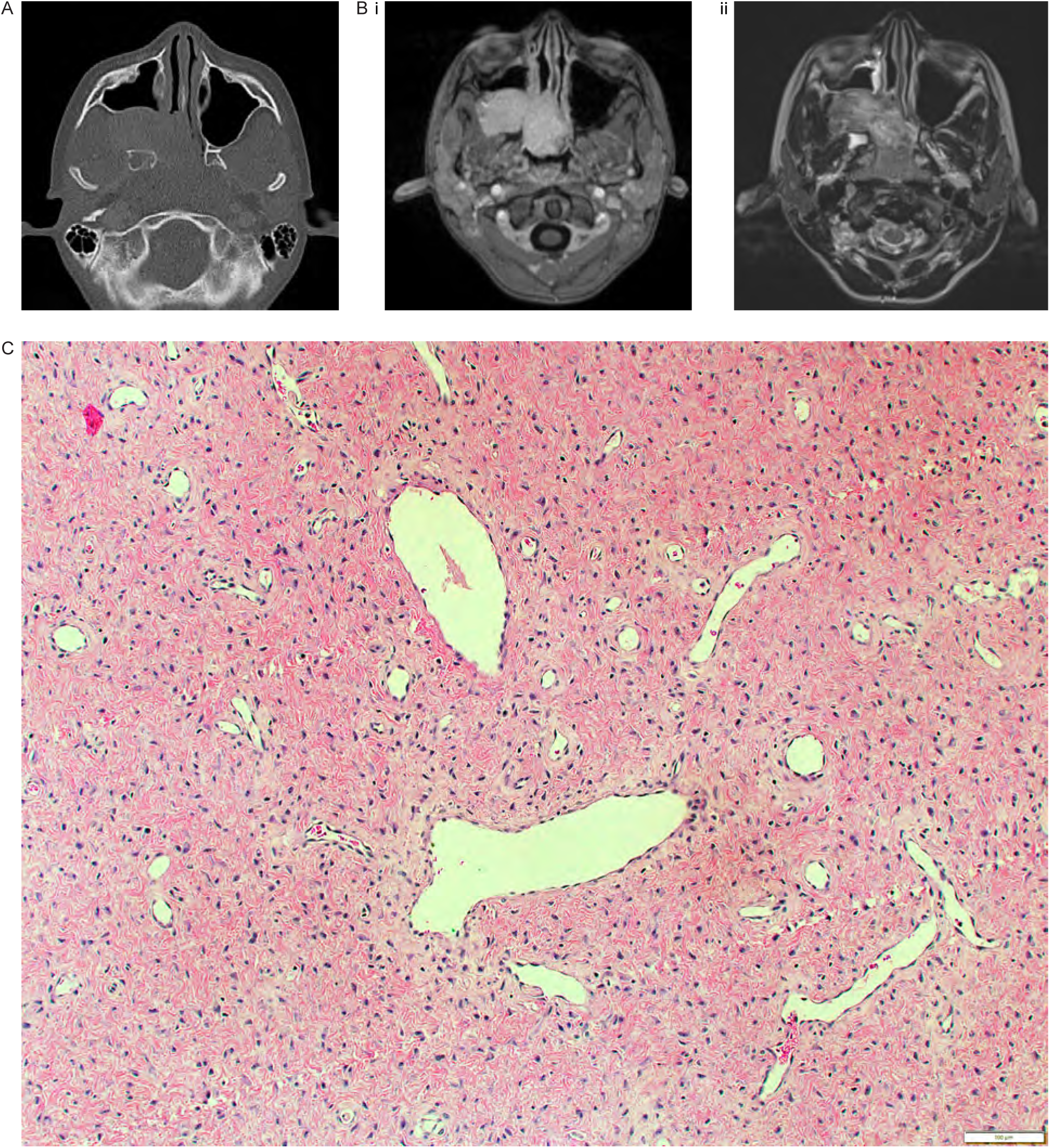
Representative radiographic and histopathologic features of juvenile nasopharyngeal angiofibroma (JNA). A Axial slice of computed tomography (CT) sinus imaging without contrast, demonstrating the Holman-Miller sign. B Axial slice of magnetic resonance imaging (MRI) with and without contrast, demonstrating marked enhancement on contrast-enhanced imaging (i) and heterogeneous appearance due to flow voids on T2 weighted imaging (ii). C Hematoxylin and eosin (H&E) staining of JNA tumor demonstrating a highly vascular fibrous stroma. Note the dilated thin-walled vessels and collagenous stroma with spindle cells (scale bar 100 µm).

Our experimental workflow is depicted in Figure 2. Briefly, samples underwent scRNA-seq, followed by cell type clustering, differential expression analysis, and computational analysis to map the tumor microenvironment and generate candidate drug-target relationships. This approach enables cell type–resolved interrogation of vascular, stromal, and immune compartments within JNA. Because JNA is defined clinically by vascularity, operative bleeding risk, and recurrence in selected patients, we focused subsequent analyses on the fibrovascular populations most likely to contribute to these disease features.

**Figure 2.**
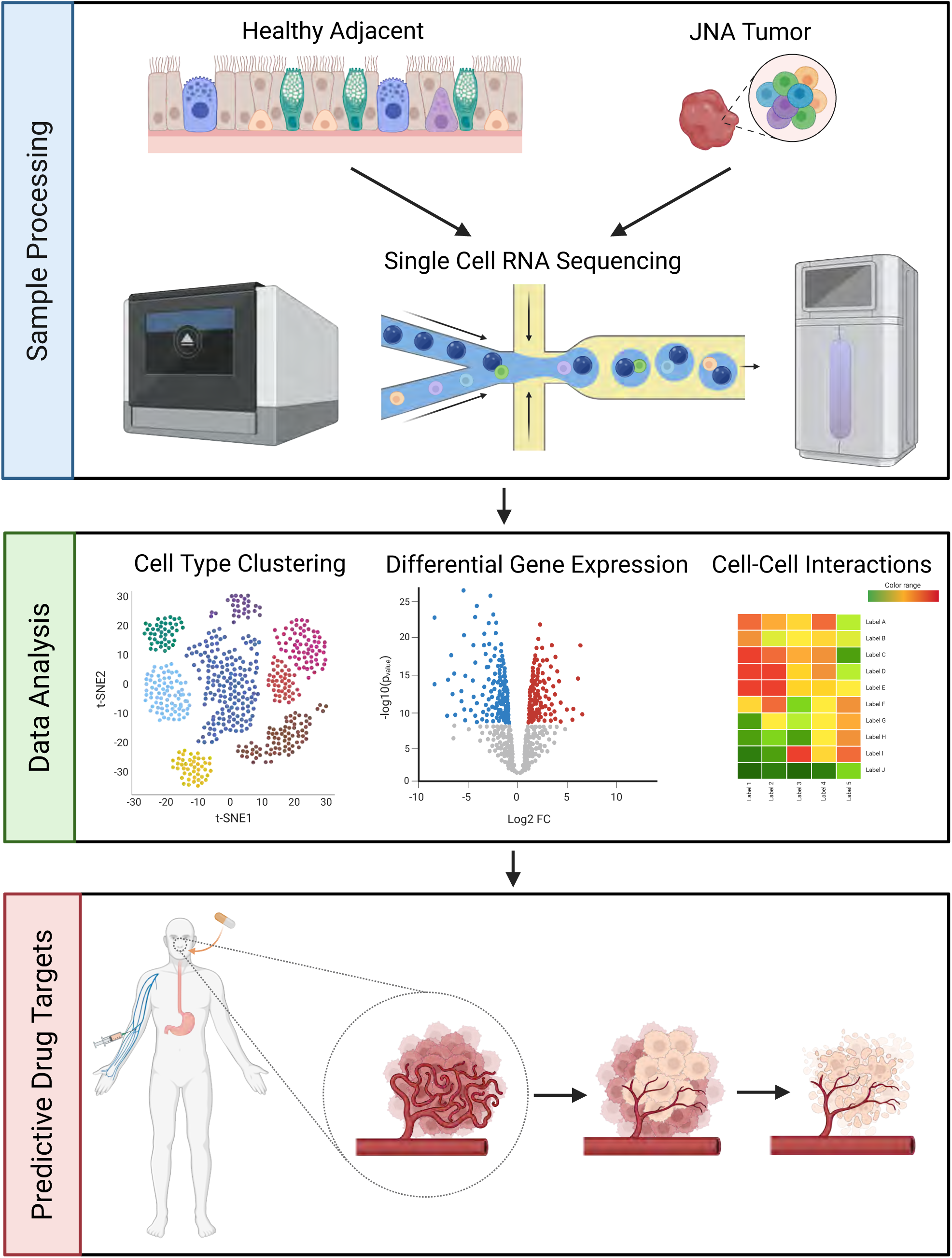
Overview of the experimental and computational workflow used to define the JNA tumor microenvironment. Tumor and tumor-adjacent sinonasal tissues were collected from patients with JNA and processed for single-cell RNA sequencing. Following sample processing and sequencing, cells were analyzed by cell-type clustering, differential gene expression, and cell-cell communication analyses to identify disease-associated cellular compartments and signaling networks. Drug-target mapping was then performed to nominate candidate therapeutically actionable pathways within the JNA fibrovascular microenvironment. Created with BioRender.com.

Samples were processed for 10x scRNA-seq analysis. A total of 19,144 cells passed quality control and were annotated into 25 major cell types based on expression of well-known cell type specific markers (Figure 3a-b). Five dominant cell types were noted, including endothelial cells, fibroblasts, pericytes, vascular smooth muscle cells, and neural crest-like cells, with more focused analysis performed on this group (Figure 3b-d). These populations correspond to the major stromal and vascular elements recognized histologically in JNA and provided a cellular framework for interrogating the mechanisms underlying tumor vascularity.

**Figure 3.**
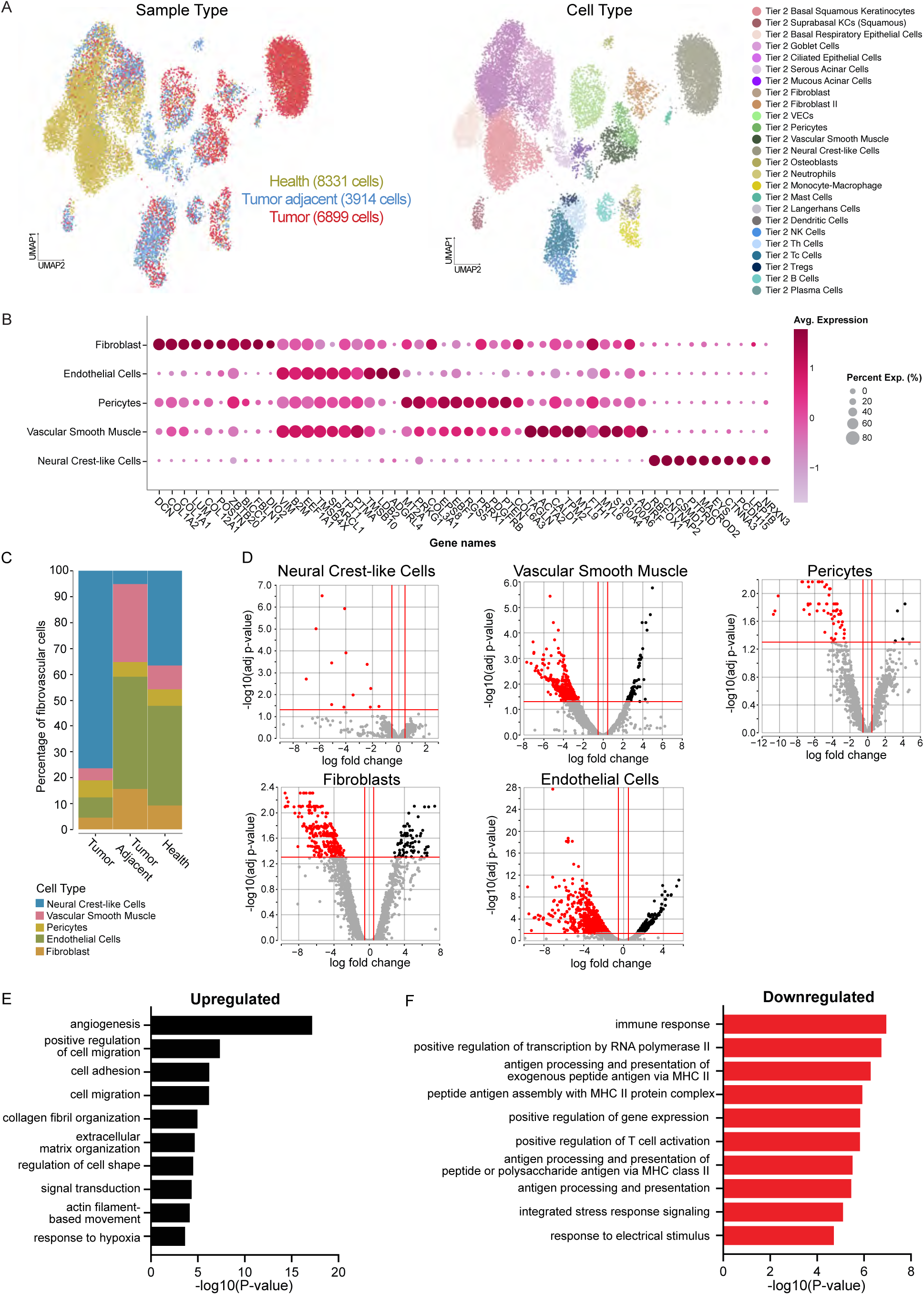
Single-cell RNA sequencing defines the cellular composition and endothelial transcriptional remodeling of JNA. A Uniform Manifold Approximation and Projection (UMAP) representation of scRNA-seq analysis of 19,144 cells from JNA (n=2), JNA adjacent (n=2), and healthy human intranasal tissue, including 6,899 tumor cells, 3,914 tumor-adjacent cells, and 8,331 healthy control cells. Cell-type annotation was based on the expression of markers shown in B. B Average scaled expression levels of selected signature genes for different cell types. C Stacked barplot showing the cell-type compositions comparing JNA, JNA adjacent, and healthy intranasal tissue. Fibrovascular populations, including endothelial cells, fibroblasts, pericytes, vascular smooth muscle cells, and neural crest-like cells, were enriched and selected for focused downstream analysis. D Volcano plots showing differentially expressed genes (DEGs) for the five predominant fibrovascular cell types in JNA compared to tumor-adjacent mucosa. Upregulated genes are highlighted in black, and downregulated genes are highlighted in red. E Gene ontology enrichment analysis of biological processes using Database for Annotation, Visualization, and Integrated Discovery (DAVID) for upregulated genes in endothelial cells. F Gene ontology enrichment analysis for downregulated genes in endothelial cells in JNA versus tumor-adjacent mucosa. Modified Fisher exact test was used to identify significantly enriched pathways (P<0.01).

### Endothelial Cells Drive an Angiogenesis-Dominant Transcriptional Program in JNA

Cell type composition analysis between JNA, tumor-adjacent tissue and healthy sinonasal tissue revealed a marked expansion of neural crest-like cells in JNA (Figure 3c). This finding is consistent with proposed developmental origins of JNA from neural crest–derived vascular structures.^9^ Despite this increase in the neural crest-like cells, very few differentially expressed genes (DEGs) were noted in this population comparing JNA to tumor-adjacent tissue (Figure 3d).

In contrast, endothelial cells had the most DEGs, with 825 genes, 234 of which were significantly upregulated (Figure 3d). This degree of transcriptional change exceeded that observed in all other major cell populations. Gene ontology analysis of the upregulated genes in endothelial cells revealed enrichment of pathways involving angiogenesis, extracellular matrix organization, and cell migration (Figure 3e). These pathways align with the clinical behavior of JNA as a highly vascular tumor with local expansion and prominent stromal remodeling. Similar analysis of the downregulated genes in endothelial cells predominantly featured pathways involving the adaptive immune system (Figure 3f). These findings support an angiogenesis-driven tumor phenotype, and this transcriptional profile suggests endothelial activation coupled with reduced expression of adaptive immune signaling within the tumor microenvironment.

Given the prominent transcriptional profile of the endothelial cell population and their potential role in the tumor microenvironment, we performed additional subclustering to further classify these cells as either lymphatic endothelial cells (LECs) or vascular endothelial cells (VECs) for the remainder of our analysis. This distinction allowed us to evaluate whether angiogenic and drug-targetable programs were concentrated within specific endothelial compartments.

### JNA Is Organized as a Fibrovascular Signaling Network

Analysis of cell-cell interactions using CellPhoneDB demonstrated increased interactions between multiple members of the fibrovascular network including fibroblasts, endothelial cells, neural crest-like cells, pericytes, and vascular smooth muscle cells in JNA (Figure 4a) versus the tumor-adjacent tissue where interactions stemmed primarily from fibroblasts to other cell types (Figure 4b). These five cell types emerge in JNA as a module of increased connectivity and signaling. This pattern suggests that JNA is not simply enriched for fibrovascular cell types but is organized around coordinated communication among stromal and vascular populations.

**Figure 4.**
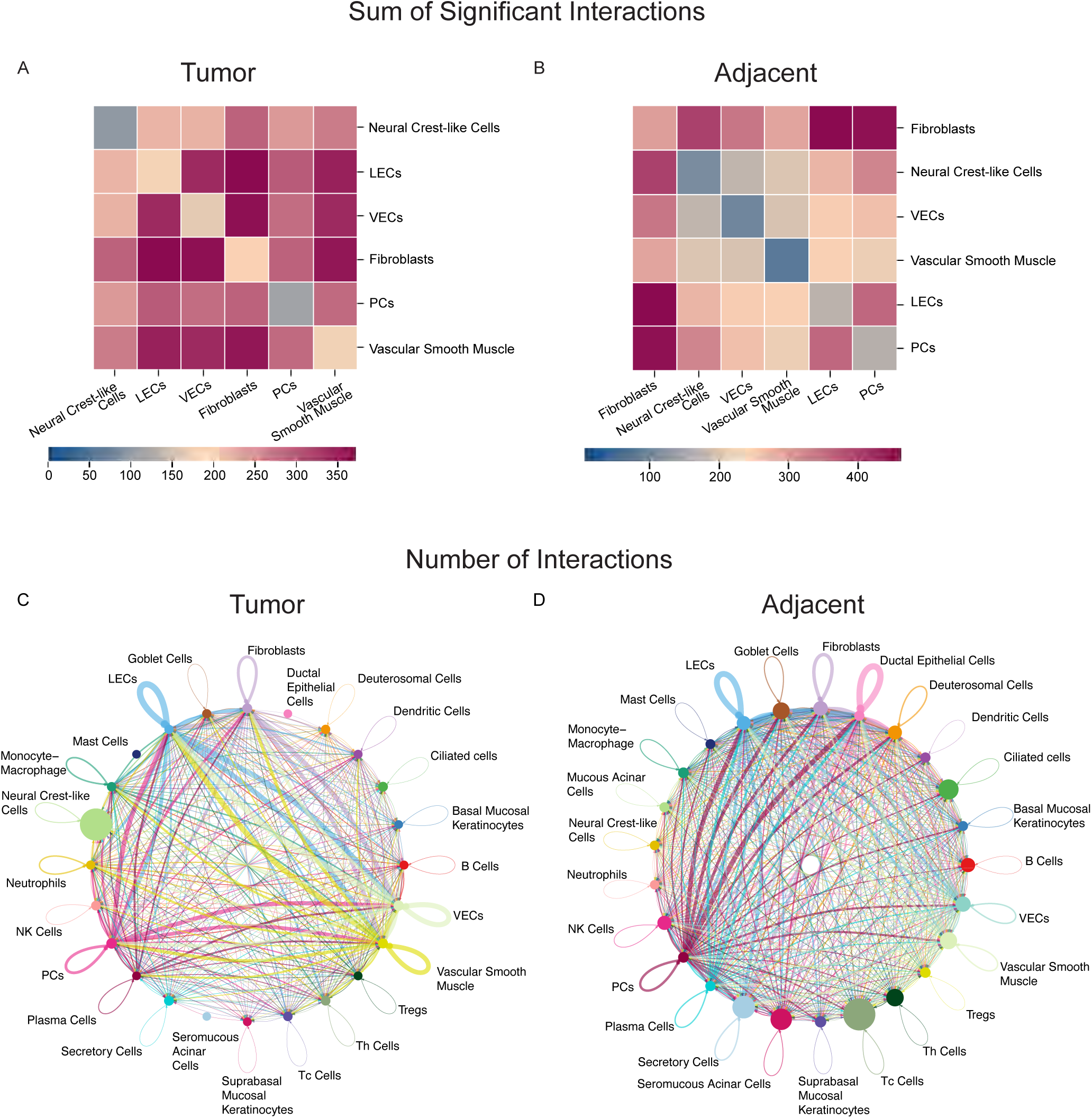
JNA is organized around a dense fibrovascular cell-cell communication network. A-B Heatmaps of CellPhoneDB analysis showing the sum of significant interactions in A JNA tumor and B tumor-adjacent mucosa. The focused fibrovascular analysis includes fibroblasts, neural crest-like cells, lymphatic endothelial cells (LECs), vascular endothelial cells (VECs), pericytes (PCs), and vascular smooth muscle cells. C-D CellChat interaction maps showing the number of interactions across all annotated cell types in C JNA tumor and D tumor-adjacent mucosa. JNA demonstrates dense preferential connectivity among LECs, VECs, vascular smooth muscle cells, pericytes, and fibroblasts, whereas tumor-adjacent mucosa shows a broader and more distributed interaction pattern across epithelial, immune, stromal, and vascular compartments.

Visualization of interaction networks via CellChat revealed dense and preferential connectivity among stromal and vascular compartments in JNA, including vascular endothelial cells, pericytes, vascular smooth muscle cells, lymphatic endothelial cells, and fibroblasts (Figure 4c), whereas healthy tissue maintained broader, less structured interactions across diverse cell populations including secretory, seromucous acinar, ciliated and ductal epithelial cells, as well as Tc cells (Figure 4d).

### Drug2cell Analysis Identifies VEGF/VEGFR Signaling as a Targetable Axis

Drug2cell analysis of JNA and tumor-adjacent tissue was performed to systematically map transcriptional programs to candidate therapeutic targets within specific cell populations.^29^ This approach leverages curated drug–gene interaction databases to infer cell type–specific therapeutic vulnerabilities directly from single-cell transcriptomic data, enabling prioritization of actionable pathways within complex tissue environments.

In JNA, this analysis revealed enrichment of drug targets associated with angiogenic signaling pathways, with a dominant focus on VEGF/VEGFR-mediated signaling (Figure 5). FDA-approved tyrosine kinase inhibitors targeting VEGFR, including lenvatinib, tivozanib, axitinib, and pazopanib, identified as candidate agents for future evaluation, with predicted activity localized predominantly to vascular endothelial and lymphatic endothelial cell populations (Figure 5). This cell type–specific enrichment supports a model in which therapeutic targeting of endothelial signaling may disrupt the broader fibrovascular network that sustains tumor growth. These findings position VEGF-driven angiogenic signaling as a dominant and targetable pathway within JNA and suggest a potential role for anti-angiogenic therapies in patients with recurrent or unresectable disease.

**Figure 5.**
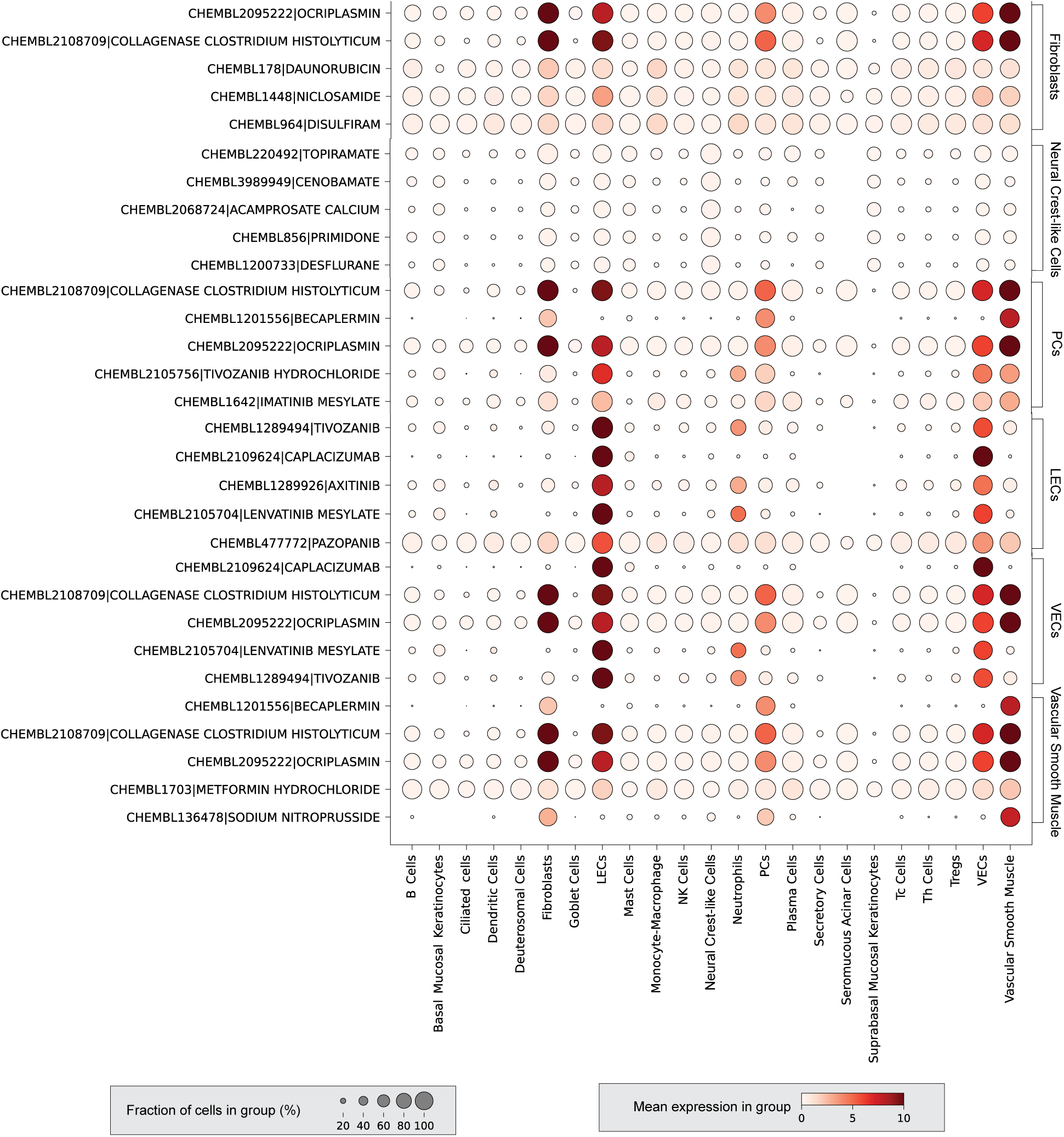
Drug2cell nominates VEGF/VEGFR-directed therapeutic vulnerabilities within the JNA fibrovascular compartment. Dot plot showing predicted Drug2cell/ChEMBL drug–gene signature enrichment across annotated cell populations in JNA. Dot size represents the fraction of cells within each group expressing the relevant drug-associated gene signature, and color intensity represents mean expression within the group. VEGFR-targeting agents, including lenvatinib, tivozanib, axitinib, and pazopanib, show enrichment predominantly within vascular endothelial cells (VECs) and lymphatic endothelial cells (LECs), supporting VEGF/VEGFR signaling as a candidate targetable axis within the JNA fibrovascular microenvironment.

## Discussion

JNA is a rare disease, which has limited the availability of comprehensive molecular datasets and hindered systematic investigation of its underlying biology. In our exploratory study, we were able to define JNA as an angiogenesis-driven fibrovascular tumor organized around endothelial-centered signaling networks through scRNA-seq. Additionally, we applied Drug2cell,^29^ a computational framework that maps single-cell transcriptional programs to candidate drug targets in a cell type–specific manner, to identify potentially actionable pathways within the tumor microenvironment, as has been shown in our previous studies.^30,31^ To the best of our knowledge, this study represents the first single-cell resolution characterization of JNA and extends emerging single-cell atlases of head and neck tissues,^32–34^ including efforts within and related to the Human Cell Atlas Oral and Craniofacial Atlas.^35^ These findings provide a cell-resolved framework for linking the classic clinical vascularity of JNA to specific stromal, endothelial, and drug-targetable programs.

Within our annotated atlas, we noted the expansion of neural crest-like cells in JNA, which is consistent with previously proposed biologic mechanism of the disease. Schick et al hypothesized that JNAs arose from the first branchial arch artery/plexus remnants and were derived from neural crest cells, based on the presence of neural crest-like cells along pathologic vessels in JNA on immunohistochemical staining.^9^ Neural crest cells are crucial in craniofacial development and play an important role in vascular development in the branchial arches, which may explain JNA’s close association with the sphenopalatine foramen and its blood supply from the internal maxillary artery as related to the first branchial arch.^36^ Despite the predominance of neural crest-like cells identified in JNA, only 13 DEGs were present in this population compared to tumor-adjacent tissue. Neural crest-like cells also engaged in fewer cell-cell interactions compared to fibroblasts, endothelial cells, vascular smooth muscle cells, pericytes, and lymphatic endothelial cells. These findings suggest that the primary active communication in JNA is among the fibrovascular components of the tumor, rather than the neural crest-like cells. While neural crest-like cells may be key players in the initial development of JNA, their active role appears to be diminished in these tumors at the time of resection. This distinction is important because it separates a potential developmental origin of JNA from the active cellular programs that may sustain tumor growth and vascular remodeling in established lesions.

In contrast, endothelial cells were noted to have the highest number of DEGs in JNA. Enriched pathways included those related to angiogenesis, extracellular matrix, and cell migration, a finding consistent with a highly vascular, proliferative tumor. Prior studies support these findings with immunohistochemical and focused transcriptomic assays demonstrating increased expression of VEGF, FGF, TGF-beta, and PDGF in JNA.^10,17,18,37,38^ Our work also demonstrates downregulation of adaptive immune pathways, specifically MHC-II related pathways, suggesting endothelial cells in JNA are in an immune inert state. Previous studies have also identified immune cells in JNA on immunohistochemical staining, primarily T-cells and mast cells,^38–41^ although, this immune infiltrate was often localized in the subepithelial compartment, associated with thrombosis and infarction.^39,40^ Endothelial cells play a crucial role in immune cell signaling and trafficking, and can even function as antigen presenting cells and recruit T-cells in inflammatory conditions.^42–44^ Such inflammatory conditions increase expression of MHC-II and other inflammatory factors.^42,45,46^ However, VEGF signaling, an important driver of angiogenesis that is upregulated in JNA, has been shown to have immunosuppressive effects in cancer, particularly with regard to T-cell activation.^47–51^ Hence, the pro-angiogenic nature of JNA may contribute to reduced adaptive immune engagement through VEGF-associated mechanisms. This could have implications for the limited role of immunomodulatory approaches in this disease, although this hypothesis requires validation in larger cohorts and protein- or spatially resolved studies.

From a therapeutic standpoint, these findings further support further evaluation of VEGF/VEGFR as possible neoadjuvant or adjuvant therapy for unresectable or recurrent JNA, respectively. This proposition is further substantiated by Drug2cell analysis, which nominated VEGFR signaling as a possible drug target in JNA. Furthermore, the nominated FDA-approved drugs that block VEGFR signaling are predicted to preferentially map the fibrovascular elements of the JNA tumor, suggesting a degree of cell-type specificity within the tumor ecosystem. These findings are consistent with the transcriptional and interaction-based analyses, such as CellPhoneDB and CellChat, identifying endothelial cells as the primary active compartment and the fibrovascular network as the central organizing structure of the tumor. Importantly, the predicted drug-target signal localized primarily to vascular endothelial and lymphatic endothelial populations, supporting the concept that the therapeutic vulnerability is linked to the same cellular compartment driving the angiogenic phenotype. The convergence of transcriptional, interaction-based, and drug-mapping analyses on VEGF signaling highlights this pathway as a coherent and biologically grounded therapeutic axis. Clinically, these data do not establish efficacy of anti-angiogenic therapy in JNA. Rather, they provide a rationale for prospective evaluation of VEGF/VEGFR-directed strategies in selected high-risk settings, including recurrent or unresectable disease.

This work serves as an important, novel, exploratory analysis of an extremely rare neoplasm; however, there are several limitations given the small sample size. Prior studies have suggested heterogeneity among JNA tumors in terms of expression patterns, which is unlikely to be fully captured in our study, thereby limiting generalizability.^37^ The cross-sectional nature of this study also limits the ability to assess temporal dynamics of tumor development. In addition, comparisons were made between tumor, tumor-adjacent, and healthy sinonasal tissues from adults undergoing minimally invasive pituitary surgery. The age differences and possibility of tumor influences on adjacent tissue could introduce confounding factors.

While single-cell RNA sequencing enables high-resolution characterization of cellular composition and transcriptional states, it does not directly assess protein expression, spatial organization, or functional activity of identified signaling pathways. As such, the inferred cell–cell interactions and pathway enrichments, including VEGF/VEGFR signaling, require validation using orthogonal approaches such as spatial transcriptomics, multiplex immunofluorescence, or functional assays.

Furthermore, while Drug2cell analysis provides a framework for identifying candidate therapeutic targets, these predictions are based on computational inference and require validation in preclinical models and prospective clinical studies. Future work should focus on expanding cohort size, incorporating spatially resolved and longitudinal datasets, and developing experimental models that recapitulate the fibrovascular architecture of JNA. Given that JNA primarily affects adolescent patients, future therapeutic studies should also carefully consider safety, timing, and risk-benefit tradeoffs for systemic anti-angiogenic therapy.

## Conclusions

In summary, scRNA-seq provides (1) a high-resolution map of the JNA tumor environment, emphasizing its neural crest-associated composition and angiogenesis-dominant fibrovascular architecture origin and (2) nominates VEGF/VEGFR signaling as a candidate therapeutic vulnerability in JNA. Additional investigation is needed to validate these findings, particularly the therapeutic implications of VEGF pathway inhibition.

## Disclosures

No authors have any financial conflicts of interest.

## Fundings Sources

This work was supported by the Chan Zuckerberg Initiative (CZI Pediatric Networks for the Human Cell Atlas, “Mapping the Pediatric Inhalation Interface”, Grant Number: 2021-237918 to AK and KMB). It was also supported by start-up funds from the ADA Science & Research Institute (Volpe Research Scholar Award), the Department of Oral and Molecular Craniofacial Biology, Philips Institute for Oral Health Research, and the VCU Massey Comprehensive Cancer Center.

## Resource availability

### Lead contact

Further information and requests for resources and reagents should be directed to and will be fulfilled by the lead contact, Kevin Matthew Byrd.

### Materials availability

This study did not generate new unique reagents. Reasonable requests for additional information regarding materials will be addressed by the lead contact.

#### Data Availability

Sequencing data generated during the study is available through GEO. Code generated during this study can be found at the following github link: https://github.com/Loci-lab/JNA_analysis

## Acknowledgements

Kevin Matthew Byrd would like to extend a heartfelt thanks to the Human Cell Atlas Consortium, specifically Sarah Teichmann, Aviv Regev, Ellen Todres, and Lucia Robson, as well as the Human Cell Atlas Oral & Craniofacial Bionetwork, for supporting this and ongoing projects to map oral and craniofacial tissues in health and disease. Furthermore, we are grateful to support from the Chan Zuckerberg Initiative for support throughout the last 5 years. This work was supported by generous start-up funds from the ADA Science & Research Institute (Volpe Research Scholar Award), the Department of Oral and Molecular Craniofacial Biology, the Philips Institute for Oral Health Research, and the VCU Massey Comprehensive Cancer Center, Massey and this work have also been supported in part by NIH-NCI Cancer Center Support Grant CA016059. This publication is part of the Human Cell Atlas initiative: http://www.humancellatlas.org/publications

## Ethical Considerations

The protocols (UNC IRB # 03-1396) were reviewed and approved by the institutional review board at the University of North Carolina. The study was conducted in compliance with the Declaration of Helsinki. All participants or their legal guardians provided written, informed consent to participate.

## Author Contributions

For this study, KMB and AJK conceptualized the project. BDT, CKC, CSE, BAS, JRV, and AJK supported the recruitment of patients and collected data. AL, KO, HM, MF, LM, and WKO performed experimental assays to generate study data. HD, MK, BTR, HMS, KMB, SHR, and AJK performed experimental and/or bioinformatic analyses that supported project development. FA performed H&E staining and provided interpretation of histopathology. HMS, BTR, and KMB wrote the original draft; HMS, BTR, AJK, SHR, FA, and KMB critically reviewed and edited the final manuscript.

## Notes

### Competing Interest Statement

The authors have declared no competing interest.

### Author Declarations

The institutional review board of the University of North Carolina - Chapel Hill gave ethical approval for this work.

## References

1. Doody J, Adil EA, Trenor CC, Cunningham MJ. The genetic and molecular determinants of juvenile nasopharyngeal angiofibroma: A systematic review. Ann Otol Rhinol Laryngol. 2019;128(11):1061–1072. doi:10.1177/0003489419850194

2. Boghani Z, Husain Q, Kanumuri VV, et al. Juvenile nasopharyngeal angiofibroma: a systematic review and comparison of endoscopic, endoscopic-assisted, and open resection in 1047 cases. Laryngoscope. 2013;123(4):859–869. doi:10.1002/lary.23843

3. López F, Triantafyllou A, Snyderman CH, et al. Nasal juvenile angiofibroma: Current perspectives with emphasis on management. Head Neck. 2017;39(5):1033–1045. doi:10.1002/hed.24696

4. Leong SC. A systematic review of surgical outcomes for advanced juvenile nasopharyngeal angiofibroma with intracranial involvement. Laryngoscope. 2013;123(5):1125–1131. doi:10.1002/lary.23760

5. Hameed N, Keshri A, Manogaran RS, et al. Intracranial extension of juvenile nasopharyngeal angiofibroma: patterns of involvement with a proposed algorithm for their management. J Neurosurg Pediatr. 2025;35(4):407–416. doi:10.3171/2024.9.PEDS24362

6. Attya HMA, Hassouna MS, Shawky AA, Abdelmalek ME. Recurrent angiofibroma: analysis of risk factors and common sites of recurrence. Eur Arch Otorhinolaryngol. June 3, 2025. doi:10.1007/s00405-025-09476-9

7. Glad H, Vainer B, Buchwald C, et al. Juvenile nasopharyngeal angiofibromas in Denmark 1981-2003: diagnosis, incidence, and treatment. Acta Otolaryngol. 2007;127(3):292–299. doi:10.1080/00016480600818138

8. Nicolai P, Berlucchi M, Tomenzoli D, et al. Endoscopic surgery for juvenile angiofibroma: when and how. Laryngoscope. 2003;113(5):775–782. doi:10.1097/00005537-200305000-00003

9. Schick B, Pillong L, Wenzel G, Wemmert S. Neural crest stem cells in juvenile angiofibromas. Int J Mol Sci. 2022;23(4). doi:10.3390/ijms23041932

10. Pandey P, Mishra A, Tripathi AM, et al. Current molecular profile of juvenile nasopharyngeal angiofibroma: First comprehensive study from India. Laryngoscope. 2017;127(3):E100–E106. doi:10.1002/lary.26250

11. Hota A, Sarkar C, Gupta SD, Kumar R, Bhalla AS, Thakar A. Expression of vascular endothelial growth factor in Juvenile Angiofibroma. Int J Pediatr Otorhinolaryngol. 2015;79(6):900–902. doi:10.1016/j.ijporl.2015.03.033

12. Brieger J, Wierzbicka M, Sokolov M, Roth Y, Szyfter W, Mann WJ. Vessel density, proliferation, and immunolocalization of vascular endothelial growth factor in juvenile nasopharyngeal angiofibromas. Arch Otolaryngol Head Neck Surg. 2004;130(6):727–731. doi:10.1001/archotol.130.6.727

13. Ponti G, Losi L, Pellacani G, et al. Wnt pathway, angiogenetic and hormonal markers in sporadic and familial adenomatous polyposis-associated juvenile nasopharyngeal angiofibromas (JNA). Appl Immunohistochem Mol Morphol. 2008;16(2):173–178. doi:10.1097/PAI.0b013e31806bee12

14. Zhang PJ, Weber R, Liang H-H, Pasha TL, LiVolsi VA. Growth factors and receptors in juvenile nasopharyngeal angiofibroma and nasal polyps: an immunohistochemical study. Arch Pathol Lab Med. 2003;127(11):1480–1484. doi:10.5858/2003-127-1480-GFARIJ

15. Jones JW, Usman S, New J, et al. Differential gene expression and pathway analysis in juvenile nasopharyngeal angiofibroma using RNA sequencing. Otolaryngol Head Neck Surg. 2018;159(3):572–575. doi:10.1177/0194599818769879

16. Schuon R, Brieger J, Heinrich UR, Roth Y, Szyfter W, Mann WJ. Immunohistochemical analysis of growth mechanisms in juvenile nasopharyngeal angiofibroma. Eur Arch Otorhinolaryngol. 2007;264(4):389–394. doi:10.1007/s00405-006-0202-z

17. Mishra A, Jaiswal R, Amita P, Mishra SC. Molecular interactions in juvenile nasopharyngeal angiofibroma: preliminary signature and relevant review. Eur Arch Otorhinolaryngol. 2019;276(1):93–100. doi:10.1007/s00405-018-5178-y

18. Mishra A, Mishra SC, Tripathi AM, Pandey A. Clinical correlation of molecular (VEGF, FGF, PDGF, c-Myc, c-Kit, Ras, p53) expression in juvenile nasopharyngeal angiofibroma. Eur Arch Otorhinolaryngol. 2018;275(11):2719–2726. doi:10.1007/s00405-018-5110-5

19. Sitenga G, Granger P, Hepola K, Aird J, Silberstein PT. The use of flutamide for the neoadjuvant treatment of juvenile nasopharyngeal angiofibroma: a review of the literature comparing results by pubertal status and tumor stage. Int J Dermatol. 2022;61(11):1346–1352. doi:10.1111/ijd.15966

20. Seront E, Van Damme A, Legrand C, et al. Preliminary results of the European multicentric phase III trial regarding sirolimus in slow-flow vascular malformations. JCI Insight. 2023;8(21). doi:10.1172/jci.insight.173095

21. Fernández KS, de Alarcon A, Adams DM, Hammill AM. Sirolimus for the treatment of juvenile nasopharyngeal angiofibroma. Pediatr Blood Cancer. 2020;67(4):e28162. doi:10.1002/pbc.28162

22. Kasai S, Akahane K, Tamai M, et al. Dose-dependent tumor regression during sirolimus therapy in an advanced juvenile nasopharyngeal angiofibroma case. Pediatr Int. 2024;66(1):e15807. doi:10.1111/ped.15807

23. Lee-Ferris RE, Okuda K, Galiger JR, et al. Prolonged airway explant culture enables study of health, disease, and viral pathogenesis. Sci Adv. 2025;11(17):eadp0451. doi:10.1126/sciadv.adp0451

24. Huang DW, Sherman BT, Lempicki RA. Systematic and integrative analysis of large gene lists using DAVID bioinformatics resources. Nat Protoc. 2009;4(1):44–57. doi:10.1038/nprot.2008.211

25. Sherman BT, Hao M, Qiu J, et al. DAVID: a web server for functional enrichment analysis and functional annotation of gene lists (2021 update). Nucleic Acids Res. 2022;50(W1):W216–W221. doi:10.1093/nar/gkac194

26. Troulé K, Petryszak R, Cakir B, et al. CellPhoneDB v5: inferring cell-cell communication from single-cell multiomics data. Nat Protoc. 2025;20(12):3412–3440. doi:10.1038/s41596-024-01137-1

27. Jin S, Plikus MV, Nie Q. CellChat for systematic analysis of cell-cell communication from single-cell transcriptomics. Nat Protoc. 2025;20(1):180–219. doi:10.1038/s41596-024-01045-4

28. Easter QT, Fernandes Matuck B, Beldorati Stark G, et al. Single-cell and spatially resolved interactomics of tooth-associated keratinocytes in periodontitis. Nat Commun. 2024;15(1):5016. doi:10.1038/s41467-024-49037-y

29. Kanemaru K, Cranley J, Muraro D, et al. Spatially resolved multiomics of human cardiac niches. Nature. 2023;619(7971):801–810. doi:10.1038/s41586-023-06311-1

30. Matuck BF, Huynh KLA, Pereira D, et al. An integrated single-cell and spatial proteotranscriptomics atlas of fibroblast-driven immunoregulation within the human adult oral cavity. Cell Press Blue. 2026;1(1). doi:10.1016/j.cpblue.2026.100007

31. Easter QT, Alvarado-Martinez Z, Kunz M, et al. Polybacterial intracellular macromolecules shape single-cell inflammatory profiles in upper airway epithelia. npj Biofilms and Microbiomes. 2025;11(1):100. doi:10.1038/s41522-025-00735-5

32. Galvani RGA, Hidalgo AAR, Biagi-Junior CA, et al. Single-cell landscape of peripheral and tumor-infiltrating immune cells in HPV-negative HNSCC. Oncoimmunology. 2026;15(1):2605741. doi:10.1080/2162402X.2025.2605741

33. Nakagawa T, Santos J, Nasamran CA, et al. Defining the relationship of salivary gland malignancies to novel cell subpopulations in human salivary glands using single nucleus RNA-sequencing. Int J Cancer. 2024;154(8):1492–1503. doi:10.1002/ijc.34790

34. Kroehling L, Chen A, Spinella A, et al. A highly resolved integrated single-cell atlas of HPV-negative head and neck cancer. Commun Med (London). 2026;6(1). doi:10.1038/s43856-026-01401-3

35. Caetano AJ, Human Cell Atlas Oral and Craniofacial Bionetwork, Sequeira I, Byrd KM. A roadmap for the human oral and craniofacial cell atlas. J Dent Res. 2022;101(11):1274–1288. doi:10.1177/00220345221110768

36. Trainor PA. Making headway: the roles of Hox genes and neural crest cells in craniofacial development. ScientificWorldJournal. 2003;3:240–264. doi:10.1100/tsw.2003.11

37. Mishra A, Pandey A, Mishra SC. Variable expression of molecular markers in juvenile nasopharyngeal angiofibroma. J Laryngol Otol. 2017;131(9):752–759. doi:10.1017/S0022215117001372

38. Zhang M, Sun X, Yu H, Hu L, Wang D. Biological distinctions between juvenile nasopharyngeal angiofibroma and vascular malformation: an immunohistochemical study. Acta Histochem. 2011;113(6):626–630. doi:10.1016/j.acthis.2010.07.003

39. Pauli J, Gundelach R, Vanelli-Rees A, et al. Juvenile nasopharyngeal angiofibroma: an immunohistochemical characterisation of the stromal cell. Pathology. 2008;40(4):396–400. doi:10.1080/00313020802035857

40. Sánchez-Romero C, Carlos R, Díaz Molina JP, Thompson LDR, de Almeida OP, Rumayor Piña A. Nasopharyngeal Angiofibroma: A Clinical, Histopathological and Immunohistochemical Study of 42 Cases with Emphasis on Stromal Features. Head Neck Pathol. 2018;12(1):52–61. doi:10.1007/s12105-017-0824-z

41. Wendler O, Schäfer R, Schick B. Mast cells and T-lymphocytes in juvenile angiofibromas. Eur Arch Otorhinolaryngol. 2007;264(7):769–775. doi:10.1007/s00405-007-0262-8

42. Pober JS, Merola J, Liu R, Manes TD. Antigen presentation by vascular cells. Front Immunol. 2017;8:1907. doi:10.3389/fimmu.2017.01907

43. Amersfoort J, Eelen G, Carmeliet P. Immunomodulation by endothelial cells - partnering up with the immune system? Nat Rev Immunol. 2022;22(9):576–588. doi:10.1038/s41577-022-00694-4

44. Fang J, Lu Y, Zheng J, et al. Exploring the crosstalk between endothelial cells, immune cells, and immune checkpoints in the tumor microenvironment: new insights and therapeutic implications. Cell Death Dis. 2023;14(9):586. doi:10.1038/s41419-023-06119-x

45. Scott NA, Zhao Y, Krishnamurthy B, Mannering SI, Kay TWH, Thomas HE. IFNγ-Induced MHC Class II Expression on Islet Endothelial Cells Is an Early Marker of Insulitis but Is Not Required for Diabetogenic CD4+ T Cell Migration. Front Immunol. 2018;9:2800. doi:10.3389/fimmu.2018.02800

46. Manes TD, Pober JS. Antigen presentation by human microvascular endothelial cells triggers ICAM-1-dependent transendothelial protrusion by, and fractalkine-dependent transendothelial migration of, effector memory CD4+ T cells. J Immunol. 2008;180(12):8386–8392. doi:10.4049/jimmunol.180.12.8386

47. Yang J, Yan J, Liu B. Targeting VEGF/VEGFR to modulate antitumor immunity. Front Immunol. 2018;9:978. doi:10.3389/fimmu.2018.00978

48. Ohm JE, Gabrilovich DI, Sempowski GD, et al. VEGF inhibits T-cell development and may contribute to tumor-induced immune suppression. Blood. 2003;101(12):4878–4886. doi:10.1182/blood-2002-07-1956

49. Gavalas NG, Tsiatas M, Tsitsilonis O, et al. VEGF directly suppresses activation of T cells from ascites secondary to ovarian cancer via VEGF receptor type 2. Br J Cancer. 2012;107(11):1869–1875. doi:10.1038/bjc.2012.468

50. Ziogas AC, Gavalas NG, Tsiatas M, et al. VEGF directly suppresses activation of T cells from ovarian cancer patients and healthy individuals via VEGF receptor Type 2. Int J Cancer. 2012;130(4):857–864. doi:10.1002/ijc.26094

51. Kaur S, Chang T, Singh SP, et al. CD47 signaling regulates the immunosuppressive activity of VEGF in T cells. J Immunol. 2014;193(8):3914–3924. doi:10.4049/jimmunol.1303116

